# COVID-19 risk perceptions of social interaction and essential activities and inequity in the United States: Results from a nationally representative survey

**DOI:** 10.1101/2021.01.30.21250705

**Authors:** Daniel J. Erchick, Alexander J. Zapf, Prativa Baral, Jeffrey Edwards, Shruti H. Mehta, Sunil S. Solomon, Dustin G. Gibson, Smisha Agarwal, Alain B. Labrique

## Abstract

**Introduction:** Severe acute respiratory syndrome coronavirus 2 (SARS-CoV-2) related diagnoses, hospitalizations, and deaths have disproportionately affected disadvantaged communities across the United States. Few studies have sought to understand how risk perceptions related to social interaction and essential activities during the COVID-19 pandemic vary by sociodemographic factors, information that could inform targeted interventions to reduce inequities in access to care and information.

**Methods:** We conducted a nationally representative online survey of 1,592 adults in the United States to understand risk perceptions related to transmission of COVID-19 for various social and essential activities. We assessed relationships for each activity, after weighting to adjust for the survey design, using bivariate comparisons and multivariable logistic regression modeling, between responses of safe and unsafe, and participant characteristics, including age, gender, race, education, income, and political affiliation.

**Results:** Half of participants were younger than 45 years (n=844, 53.0%), female (n=800, 50.3%), and White/Caucasian (n=685, 43.0%), Black/African American (n=410, 25.8%), or Hispanic/Latino (n=382, 24.0%). Risk perceptions of unsafe for 13 activities ranged from 29.2% to 73.5%. Large gatherings, indoor dining, and visits with elderly relatives had the highest proportion of unsafe responses (>58%) while activities outdoor, visiting the doctor or dentist, and going to the grocery store had the lowest (<36%). Older respondents were more likely to view social gatherings and indoor activities as unsafe, yet more likely to view activities such as going to the grocery store, participating in outdoor activities, visiting elderly relatives, and visiting the doctor or emergency room as safe. Compared to White/Caucasian respondents, Black/African American and Hispanic/Latino respondents were more likely to view activities such as dining and visiting friends outdoor as unsafe. Generally, men vs. women, Republicans vs. Democrats and independents, and individuals with higher vs. lower income were more likely to view activities as safe.

**Conclusions:** These findings suggest the importance of sociodemographic differences in risk perception, health behaviors, and access to information and health care when implementing efforts to control the COVID-19 pandemic. Further research should address how evidence-based interventions can be tailored considering these differences with a goal of increased health equity in the pandemic response.

## INTRODUCTION

As of January 2021, severe acute respiratory syndrome coronavirus 2 (SARS-CoV-2), the virus that causes coronavirus disease (COVID-19), has infected more than 23 million people and contributed to over 390,000 deaths in the United States.^1^ The negative health and social consequences of the COVID-19 pandemic – including morbidity and mortality; decreased access to health care; and lost jobs and economic hardships – have not been experienced equally, and instead have impacted certain communities in greater numbers and with increased severity. For example, COVID-19 related diagnoses, hospitalizations, and deaths have disproportionately affected Black communities^2^ and those in poverty,^3^ demonstrating the impact of structural racism and health disparities in disadvantaged populations.^4^

Numerous COVID-19 pandemic, tracking, mapping, and monitoring tools have emerged, covering a wide array of indicators from testing capacity to daily case counts and deaths to policy interventions.^5,6^ While data collected from these trackers provide critical insights into the COVID-19 pandemic trajectory and public health response measures, they rarely address upstream socio-behavioral aspects, such as risk perceptions, knowledge and access to information, spread of misinformation, and agency and stigma. Yet access to information and health literacy vary by age, gender, and race and other characteristics with important implications for risk perceptions, behaviors, and health outcomes, including COVID-19 infection and mortality.^7^

Few studies have sought to estimate prevalence of risk perceptions related to social interaction or essential activities during the COVID-19 pandemic or explore associations between these perceptions and sociodemographic factors.^8,9^ Differences in risk perceptions could provide insights into the determinants of risk perception and health knowledge and subsequent behaviors related to the COVID-19 pandemic, while also helping to inform development of targeted communication campaigns and preventive interventions.^10,11^

The National Pandemic Pulse is a United States-population representative, internet phone/computer survey designed to obtain data on preventive behaviors, risk perceptions, agency and stigma, and misinformation related to the ongoing COVID-19 pandemic across census regions.^12^ Our aim is to examine relationships between these issues and sociodemographic factors to understand how systematic racism and inequity impact health and wellbeing in the context of the COVID-19 pandemic. Here we present findings from the first national Pandemic Pulse Survey to understand racial and sociodemographic differences in risk perceptions of social interaction and essential activities during the COVID-19 pandemic.

## METHODS

### Study population

We conducted a cross-sectional online survey of adults currently living in the U.S. ages 18 and older from September 1^st^ to 7^th^, 2020. The sample was selected from an online panel to represent the U.S. Census population using pre-specified demographic quotas for age, gender, race, census region, and income. Black/African American and Hispanic/Latino respondents were over-sampled by approximately 385 individuals per group to increase power for analyses comparing risk perceptions by ethnicity/race groups. This sample allowed for detection of a 10% difference in proportions between White, Black, and Hispanic ethnicity/race groups assuming power of 80%, type I error rate of 0.05, and a baseline prevalence of 40%-60%. Dynata – a market research firm (https://www.dynata.com) that maintains a large first-party global data platform, including 62 million panelists with accompanying demographic information – selected a random sample from their database to match the U.S. Census estimates. Dynata sent invitations by email to 16,904 panelists matching the required demographic targets of the survey until each quota was filled. The survey response rate was 10.0% and completion rate among eligible respondents was 95.3%. Survey responses were excluded for the following reasons: age less than 18 (n=47), residence outside United States (n=3), ethnicity/race for which sample quota was already filled (n=171), refusal of consent (n=72), and partial interview (n=77). Security and data quality checks utilized included digital fingerprinting and spot-checking via third-party verification to confirm the identity of the respondents and prevent duplication. Participants received a small compensation for survey completion.

### Questionnaire

A team of experts at Johns Hopkins Bloomberg School of Public Health collated COVID-19 questions from existing surveys and created new questions to address existing gaps in the literature. In a module on risk perception, the focus of this analysis, participants were presented with a series of thirteen activities related to social and essential activities and asked to respond to the question: “How safe or unsafe do you think the following activities are in terms of your getting COVID-19 or giving it to someone else?” Allowed responses included extremely safe, somewhat safe, somewhat unsafe, extremely unsafe, unsure, and prefer not to say. For the purpose of this analysis, we collapsed extremely and somewhat categories into perceptions of ‘safe’ and ‘unsafe’.

### Statistical analysis

All analyses were adjusted for the study design using survey weights for race by Census region generated using the 2010 U.S. Census estimates. We assessed bivariate relationships between responses of safe, unsafe, and unsure and participant characteristics for each activity presenting percent change (absolute) and assessing significance using Pearson’s chi-squared tests. We used multivariable logistic regression models to calculate unadjusted and adjusted odds ratios (OR and aOR) of perceiving each activity as unsafe and associated 95% confidence intervals (CIs).

Participant demographic and socioeconomic characteristics included in multivariable models were age, gender, race/ethnicity, education, income, census region, and political affiliation. To assess differences in risk perceptions by age and race, we presented relationships overall and stratified by White/Caucasian, Black/African American, and Hispanic/Latino groups.

Multivariable logistic regression models were also extended to include interaction terms for age and race and assessed for significance using Wald tests (p<0.05). Statistical analyses were conducted in Stata 16.1 (StataCorp, College Station, Texas, USA).

### Ethical approval

Participants provided electronic consent to participate by responding to a question on the survey. The study received ethical approval from the Institutional Review Board at Johns Hopkins Bloomberg School of Public Health, Baltimore, USA.

## RESULTS

### Participant characteristics

Complete responses from 1,592 respondents were included in this analysis. Roughly half of respondents were less than 45 years old (52.2%) and female (49.5%) (weighted percentages; Table 1). Participants were 60.0% White/Caucasian, 12.4% Black/African American, and 18.4% Hispanic/Latino. Risk perceptions of unsafe for the 13 activities ranged from 29.6% to 73.5% and unsure from 3.7% to 11.6% (Figure 1). Large gatherings (of 10, 100, and church), indoor dining, and visits with elderly relatives had the highest proportion of unsafe responses (>58%) while activities outdoor (dining, visiting friends), visiting the doctor or dentist, and going to the grocery store had the lowest (<36%).

**Table 1:** Participant characteristics∼.

| **Characteristic** | **n=1,592\*** | **Percent<sup>+</sup>** |
| --- | --- | --- |
| **Age (years)** |  |  |
| 18-24 | 187 | 10.3 |
| 25-34 | 352 | 21.7 |
| 35-44 | 305 | 20.2 |
| 45-54 | 245 | 16.3 |
| 55-64 | 239 | 14.7 |
| 65+ | 264 | 16.8 |
| **Gender** |  |  |
| Female | 800 | 49.5 |
| Male | 786 | 50.5 |
| Other | 1 | 0.0 |
| **Race** |  |  |
| White/Caucasian | 685 | 60.0 |
| Black/African American | 410 | 12.4 |
| Hispanic/Latino | 382 | 18.4 |
| Asian/Pacific Islander | 61 | 5.8 |
| American Indian/Alaska Native | 20 | 0.7 |
| Other | 34 | 2.8 |
| **Education** |  |  |
| High school or less | 345 | 20.2 |
| Associate degree | 215 | 13.2 |
| Some college (no degree) | 289 | 17.9 |
| Bachelor's Degree | 450 | 28.9 |
| Graduate Degree | 288 | 19.7 |
| **Income** |  |  |
| <\$20,000 | 273 | 16.3 |
| \$20,000-<\$40,000 | 317 | 19.0 |
| \$40,000-<\$70,000 | 416 | 26.9 |
| \$70,000-<\$100,000 | 258 | 16.8 |
| ≥\$100,000 | 315 | 21.0 |
| **Lost job** |  |  |
| No | 1008 | 65.3 |
| Yes | 333 | 19.8 |
| Retired | 234 | 14.9 |
| **Census region** |  |  |
| Northeast | 312 | 17.1 |
| Midwest | 347 | 20.8 |
| South | 561 | 38.3 |
| West | 372 | 23.9 |
| **Political party** |  |  |
| Republican | 429 | 39.1 |
| Democrat | 699 | 32.2 |
| Independent | 371 | 25.2 |
| Other | 52 | 3.5 |

\* Actual numbers of individuals surveyed
+ Overall population percentage adjusted for survey sample design by weighting for race by Census region.
~ Participant responses not listed above include the following “other” and “prefer not to say” categories (number, percentage adjusted for survey sample design): age: n=0; gender: refuse (n=5, 0.3%); race: n=0; education: refuse (n=5, 0.2%), income: refuse (n=13, 0.6%); lost job: refuse (n=17, 0.9%); census: n=0; and political affiliation: refuse (n=41, 2.1%).

**Figure 1:**
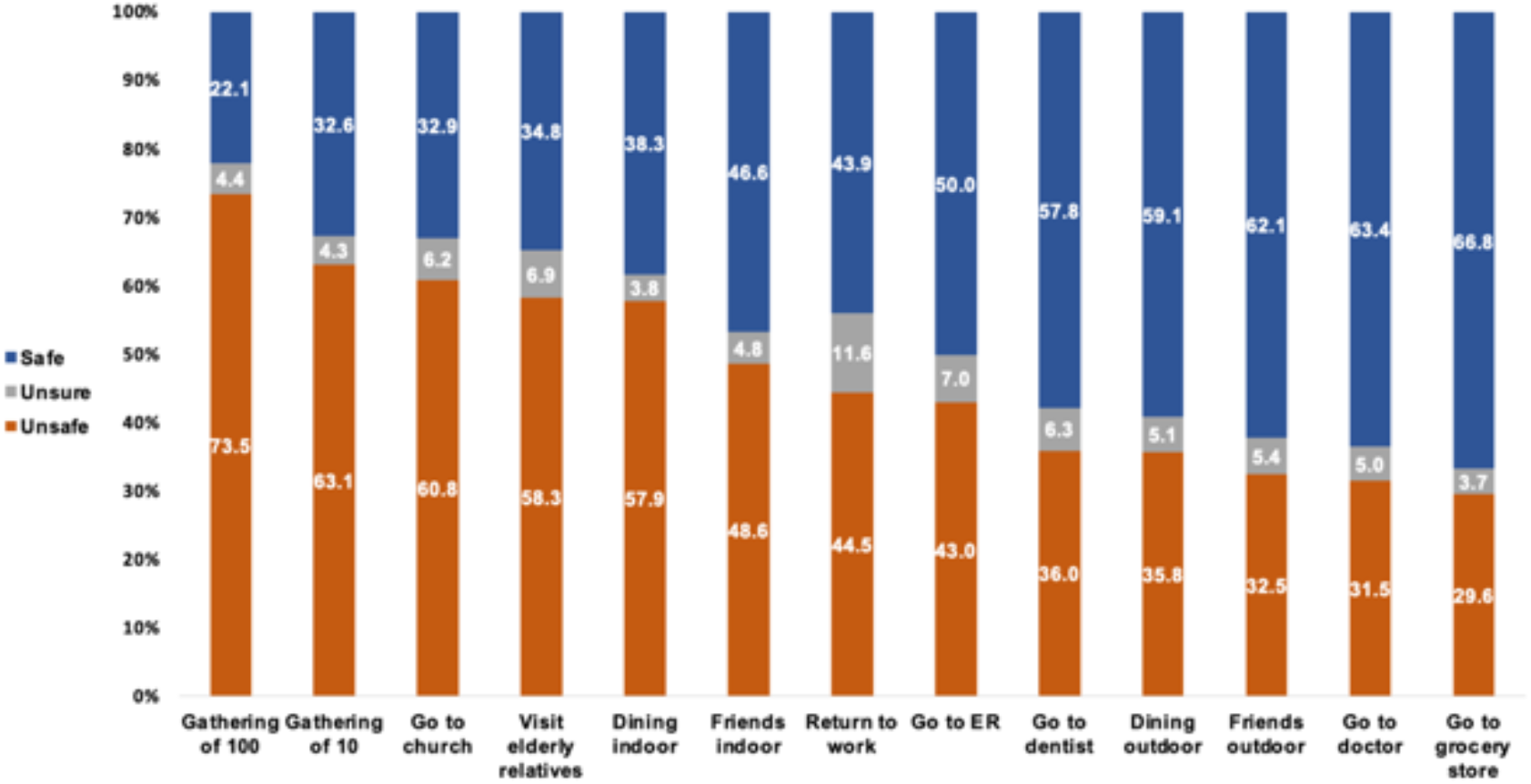
Participant risk perceptions for each activity. Percentages are the weighted estimates adjusted for race by Census region to match the overall U.S. population. Extremely safe and somewhat safe and extremely unsafe and somewhat unsafe response categories were collapsed into safe and unsafe, respectively.

### Large gatherings and activities in public

Perceptions of unsafe increased by >15% from the lowest to highest age categories for gathering of 10, gathering of 100, and going to church (all p<0.001), but decreased by a similar amount for going to the grocery store (p=0.015) (Figure 2). Males were less likely to perceive these activities as unsafe, with significant differences (p<0.05), ranging from −3.3% to 7.4%, except gathering of 10. Perceptions differed by race only for gatherings of 10, highest among Hispanic/Latino (67.5%) and Asian/Pacific Islander respondents (67.1%) (p=0.011).

**Figure 2:**
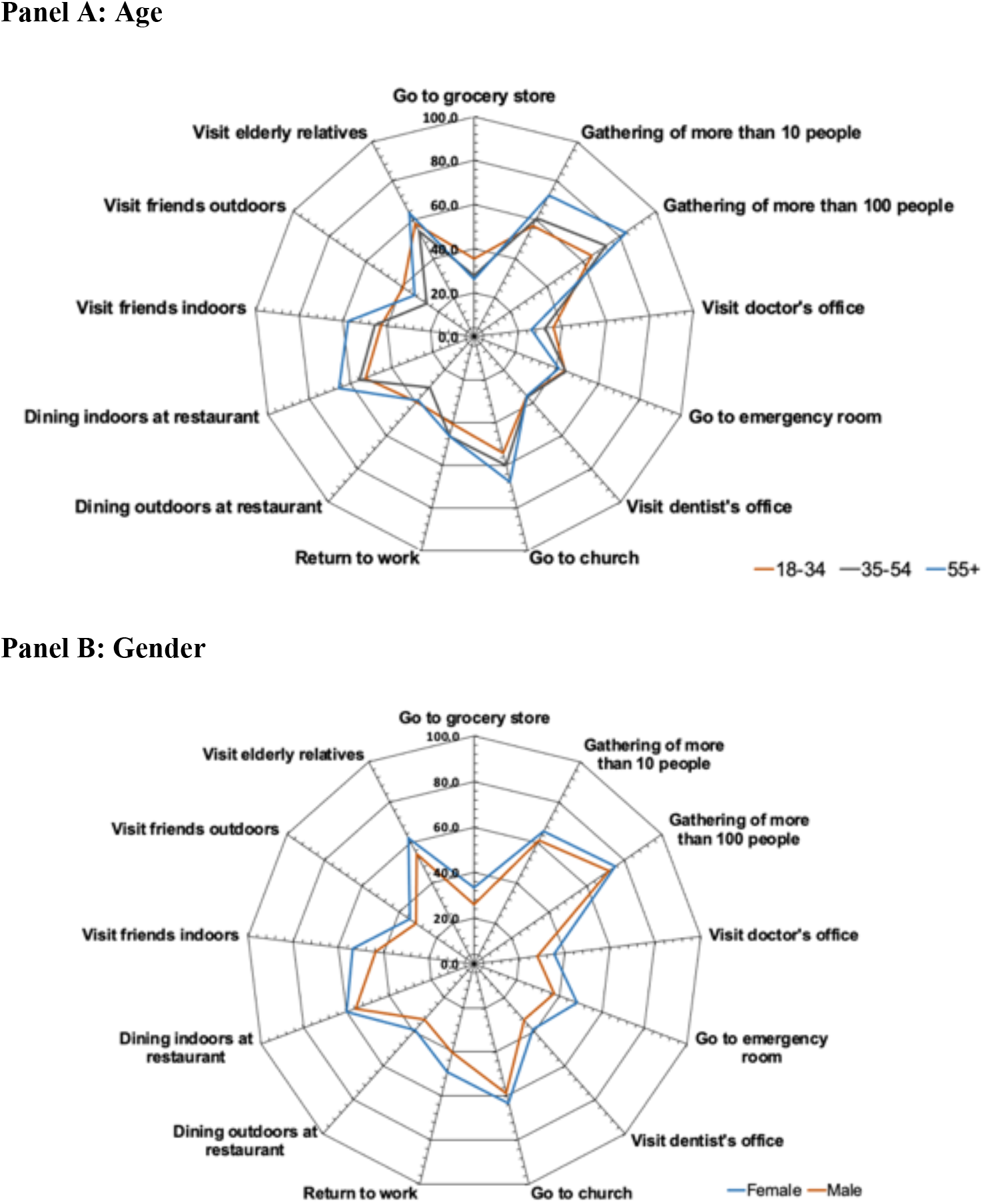
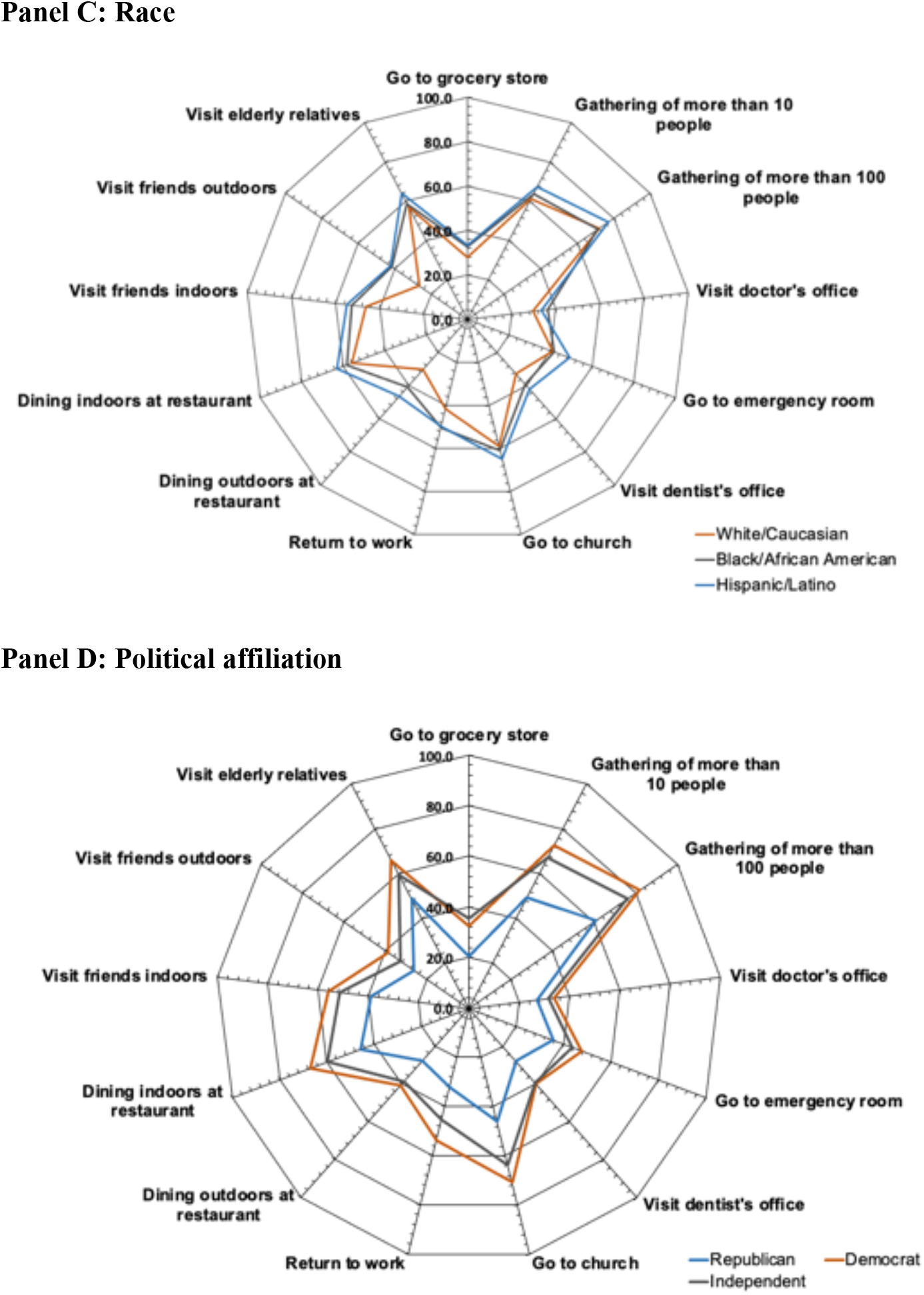
Prevalence of risk perceptions of unsafe by age, gender, race, and political affiliations. Prevalence of a perception of unsafe for each category weighted for race by Census region to match the overall U.S. population.

Respondents with high education were less likely to perceive gathering of 100 as unsafe (p=0.024). Perceptions of unsafe decreased with increasing income (p<0.05), with differences between <$20,000 and ≥$100,000 categories ranging from −3.2% to −10.2%. Democrats and independents were more likely to perceive activities as unsafe for all variables compared to Republicans (p<0.001).

In multivariable models (Figure 3 and Supplementary Table 1) perception of unsafe increased with age for gathering of 10 (aOR=1.24 (95% CI: 1.14, 1.35)), gathering of 100 (aOR=1.38 (95% CI: 1.25, 1.52)), and going to church (aOR=1.18 (95% CI: 1.09, 1.28)) and decreased for going to the grocery store (aOR= 0.89 (95% CI: 0.82, 0.96)). Men were less likely to perceive activities as unsafe. Across income groups, there was a significant decrease in perception of unsafe with increasing income for gathering of 10 (aOR=0.86 (95% CI: 0.77, 0.96)) and going to the grocery store (aOR=0.83 (95% CI: 0.74, 0.92)). Democrats and independents were more likely to report activities as unsafe relative to Republicans.

**Figure 3:**
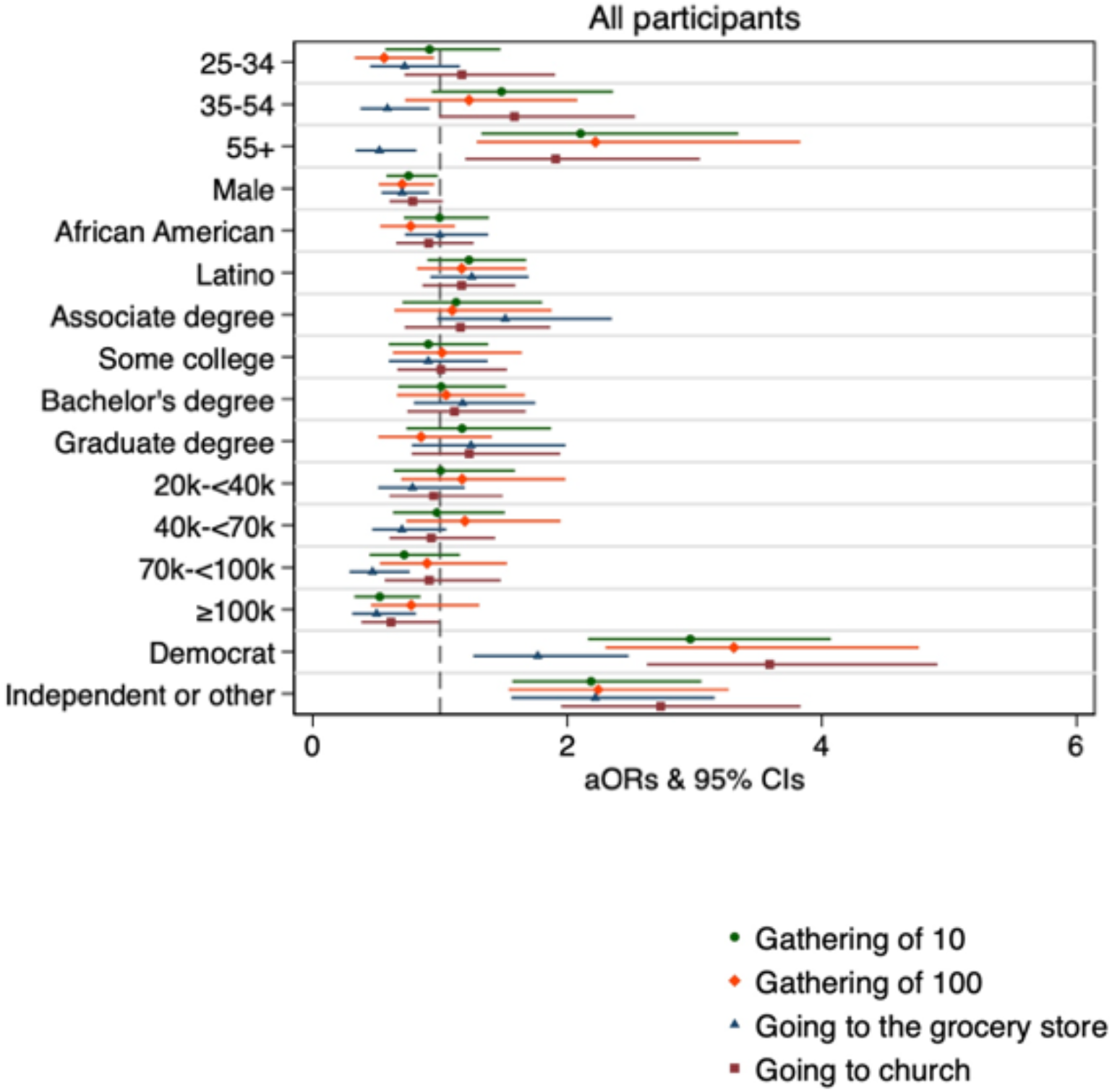
Adjusted odds ratios of perceiving large gatherings and activities in public as unsafe for all participants. Reference groups are age: 18-24, gender: female, race: White/Caucasian, education: high school or less, income: <$20,000, political party: republican.

### Indoor and outdoor dining and visits with relatives

Perceptions of unsafe increased between lowest and highest age categories by >10% for dining indoor (p<0.001) and visiting friends indoor (p=0.001), and decreased, ranging from −3.1% to − 10.1%, for visiting elderly relatives (p=0.039), visiting friends outdoor (p=0.001), and dining outdoor (p=0.006). Men were less likely to perceive activities as unsafe, with significant differences (p<0.05), ranging from −3.3% to −10.3%, except for visiting friends outdoor. Activities in this category varied by race, with White/Caucasian respondents generally less likely to perceive them as unsafe. Respondents with higher education were less likely to perceive dining outdoor as unsafe (p=0.040). Perceptions of unsafe decreased with increasing income (p<0.05) for most of these activities, ranging from −3.8% to −11.8%, except for visiting friends indoor. Democrats and independents were more likely to report activities as unsafe relative to Republicans (p<0.001).

In multivariable models (Figure 4), risk perception across age groups increased significantly for dining indoor (aOR=1.12 (95% CI: 1.04, 1.21)) and visiting friends indoor (aOR=1.15 (95% CI: 1.07, 1.24)). Men relative to women had lower odds of viewing these activities as unsafe, but this was only significant for visiting friends indoor. There was a significant decreasing trend across income groups for dining indoor (aOR=0.87 (95% CI: 0.78, 0.97)) and dining outdoor (aOR=0.87 (95% CI: 0.78, 0.96)) but not visiting friends in either setting. Compared to White/Caucasian respondents, Black/African American and Hispanic/Latino respondents were more likely to view dining outdoor and visiting friends outdoor as unsafe (Supplementary Figure 1). Democrats were more likely to view these activities as unsafe relative to Republicans. There was a statistically significant interaction between age and race for visiting an elderly relative (p=0.061) (Supplementary Table 2). The change in odds of perceiving visiting an elderly relative as unsafe for each 10-year increase in age was non-significant among White/Caucasian respondents (aOR=0.99 (95% CI: 0.89, 1.10)) and Hispanic/Latino respondents (aOR=1.11 (95% CI: 0.96, 1.29)) but significant among Black/African American respondents (aOR=1.35 (95% CI: 1.15, 1.58)).

**Figure 4:**
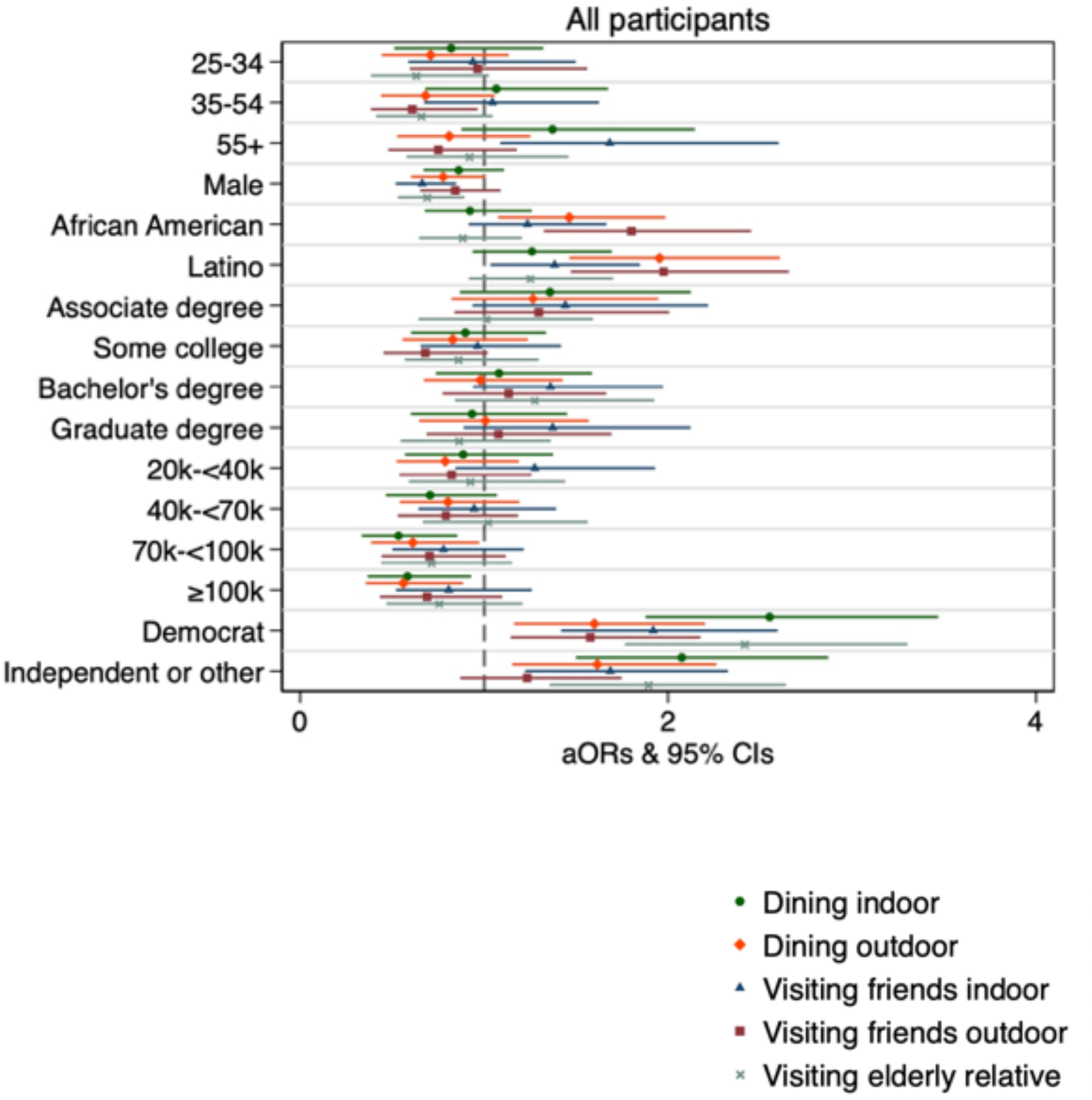
Adjusted odds ratios and 95% CIs of perceiving indoor and outdoor dining and visits with friends and relatives as unsafe for all participants. Reference groups are age: 18-24, gender: female, race: White/Caucasian, education: high school or less, income: <$20,000, political party: republican.

### Medical visits and returning to work

Perceptions of unsafe decreased (−16.2% and −6.3%, respectively) between the lowest and highest age categories for doctor visits (p<0.001) and going to the emergency room (p=0.006), and increased (4.2%) for returning to work (p<0.001). Men were less likely to perceive these activities as unsafe, with significant differences (p<0.05) ranging from −5.9% to −10.5%. Dentist visits were the only activity for which risk perception significantly differed by race (p<0.001). Respondents with lower education were more likely to respond “unsure” relative to those with higher education, with differences (p<0.05) between lowest and highest categories ranging from −5.2% to −6.9%. Respondents with higher income were less likely to perceive these activities as unsafe with a range of difference between the lowest and highest categories of −4.3% and −12.5% (p<0.05). Democrats and independents were more likely to report activities as unsafe relative to Republicans (p<0.001).

In multivariable models (Figure 5), a risk perception of unsafe across age groups decreased significantly for going to the doctor (aOR=0.84 (95% CI: 0.78, 0.91)) and emergency room (aOR=0.90 (95% CI: 0.84, 0.97)). Males were less likely to view going to the doctor, emergency room, and returning to work as unsafe. Compared to White/Caucasian respondents, Hispanic/Latino respondents were more likely to view going to the dentist or emergency room as unsafe. Respondents with higher income were less likely to view these activities as unsafe; trends across income groups were statistically significant for going to the doctor (aOR=0.84 (95% CI: 0.75, 0.94)), dentist (aOR=0.87 (95% CI: 0.78, 0.97)), and emergency room (aOR=0.86 (95% CI: 0.78, 0.96)). Democrats and independents were more likely to view activities as unsafe. There was a statistically significant interaction between age and race for returning to work (p=0.039). The change in odds of perceiving returning to work as unsafe for each 10-year increase in age was smallest for White/Caucasian respondents (aOR=1.13 (95% CI: 1.00, 1.27)) followed by Hispanic/Latino respondents (aOR=1.21 (95% CI: 1.03, 1.42)) and Black/African American respondents (aOR=1.31 (95% CI: 1.12, 1.52)).

**Figure 5:**
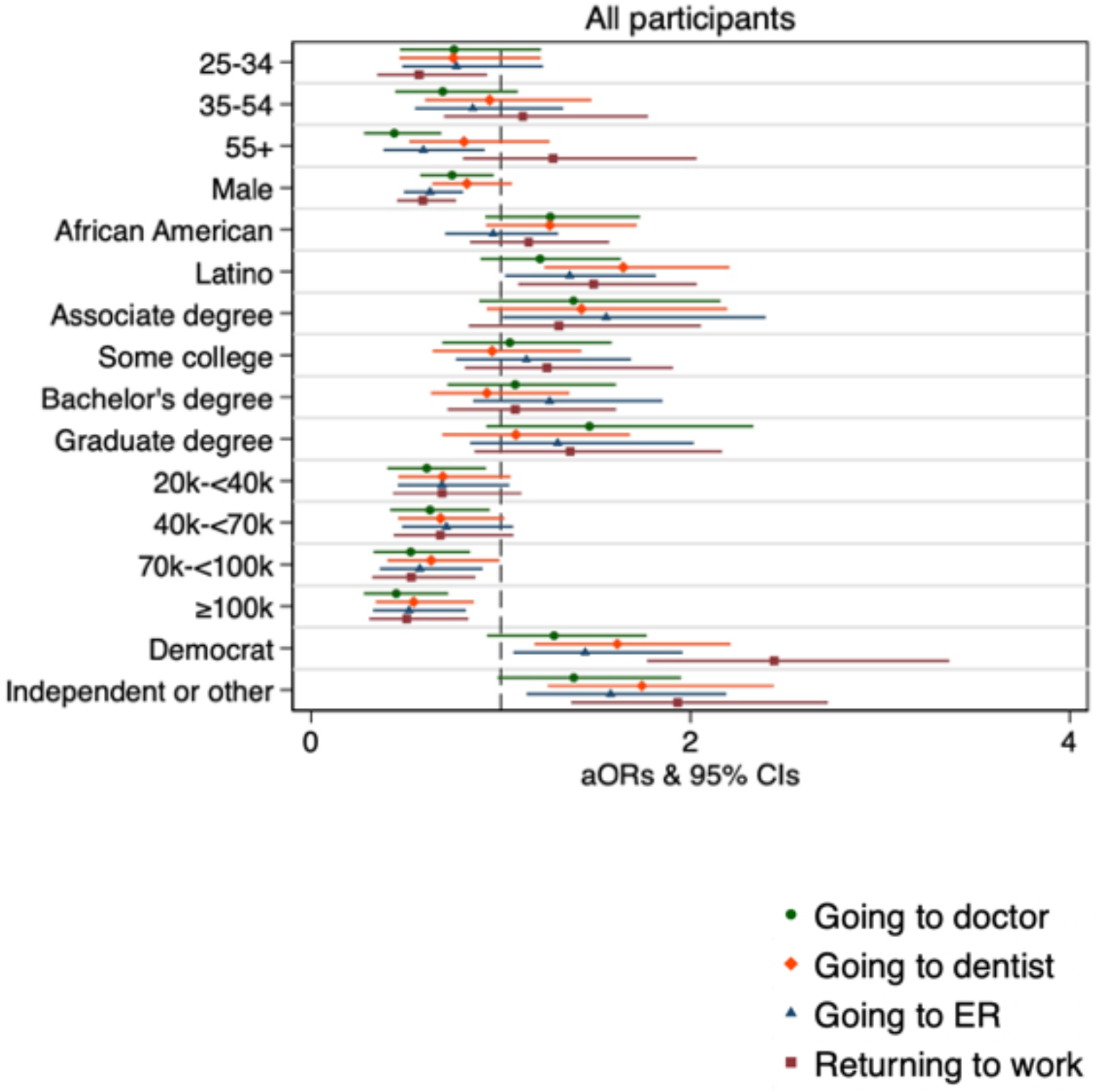
Adjusted odds ratios and 95% CIs of perceiving medical visits and returning to work as unsafe for all participants. Reference groups are age: 18-24, gender: female, race: White/Caucasian, education: high school or less, income: <$20,000, political party: republican.

### Census region

Differences between census regions in bivariate comparisons included higher proportions of respondents considering activities as unsafe in the west vs. north (gathering of 10, gathering of 100, grocery store, church, and dentist) and south vs. north (dining indoor). Census region was only predictive of risk perception in multivariable models for three activities (dining indoor: Midwest vs. Northeast: aOR=0.66 (95% CI: 0.44, 0.98); visiting friends indoor: Midwest vs. Northeast: aOR=0.68 (95% CI: 0.46, 1.00); and dining outdoor: South vs. Northeast aOR=1.44 (95% CI: 1.01, 2.06)).

## DISCUSSION

We conducted a nationally representative survey of the U.S. population to understand risk perceptions related to transmission of COVID-19 for social interaction and essential activities. Overall, risk perceptions ranged widely, but were higher for activities which have been shown to present increased risk for COVID-19 infection, particularly large gatherings and indoor activities, suggesting effective information dissemination to the public risk regarding COVID-19 risk factors.^13^ Our results suggest that risk perceptions for age and race vary by the type of activity. Men were more likely to view activities as safe compared to women, a similar finding to a large survey in eight countries that found that women were more likely to perceive COVID-19 as a serious health problem and agree and comply with restrictive public policy interventions.^14^ Individuals with higher income in our survey were more likely to view activities as safe, perhaps a result of facing fewer barriers to physical distancing.^15^ This could also reflect wealth differentials in the experience of the pandemic, with increased COVID-19 transmission and case volumes in low-income and minority populations.^16^ There were few differences by education.

Nearly universally, Democrats and independents were more likely than Republicans to view activities as unsafe, potentially a reflection of the highly polarized U.S. climate in which information about COVID-19 has been influenced by politics.

Previous studies about perceived health and economic risks associated with COVID-19 have shown significant differences in risk perception by age, gender, education, and other sociodemographic factors. A cross-sectional survey of U.S. adults conducted in March 2020 found lower risk perceptions, but higher prevalence of social distancing behaviors, among older adults.^17^ Other studies have shown mixed results by age, with some reporting higher risk perceptions for older adults^18^ and others lower.^19^ Our study showed that older respondents were more likely to view social gatherings with many people and indoor activities as unsafe, yet more likely to view activities such as going to the grocery store, participating in outdoor activities, visiting elderly relatives, and visiting the doctor or emergency room as safe.

Studies have found lower perceived risk of COVID-19 infection and mortality among Black/African American persons.^17^ Another study reported higher risk perceptions concerning COVID-19 in Native American/Alaska Native and Asian groups relative to Black/African American persons.^18^ Associations between respondent race/ethnicity and risk perceptions in our study varied by activity; for some, such as attending gatherings, visiting grocery stores, and attending church, there were no significant differences between groups. However, Black/African American and, especially, Hispanic/Latino respondents were more likely to view several activities, such as dining and visiting friends outdoor, as unsafe compared to White/Caucasian respondents. Evidence suggests that Black and Hispanic groups have higher rates of infection and mortality from COVID-19.^20^ This raises questions as to how structural racism and socioeconomic and health disparities influence access to information and trust in health services and authorities in the context of the COVID-19 pandemic. Authors of a qualitative study in a rural Latino community suggested that risk perceptions and concerns were linked to stress of loss of employment.^21^ Responsibility rests with politicians, health authorities, and community leaders to communicate evidence-based information in a manner that is honest and clear, easily accessible, and culturally appropriate. Respondents in the study of perceptions in the rural Latino community suggested, for example, a personalized approach to deliver information, by utilizing email or text messages from nearby universities, their medical providers, or the local health department.^9,21^

Perceived risks of COVID-19 morbidity and mortality have not necessarily aligned with actual behaviors.^17^ While some studies have shown close correlation between perceived disease severity and preventive behaviors, others studies have reported discrepancies between perceived disease risk and adherence to prevention behaviors; this suggests that efforts to change risk perceptions alone may be inappropriate and inadequate.^22,23^ Examining how sociodemographic factors influence risk perceptions and behaviors could identify how inequities lead to increased health risks in specific disadvantaged groups. Further, risk perceptions are likely to vary by location, local COVID-19 incidence, and over time as more information becomes available, factors such as ‘pandemic fatigue’ increase in prevalence, and more recent experiences exert a stronger influence on how people view the pandemic. In the U.S., many published studies to date were conducted during the early phases of the pandemic and focused on perceived risks of infection or mortality and health behaviors, often without detailed information on race/ethnicity.^22,24^ Our findings supplement this body of evidence by providing insights into perceived risks for specific activities, sufficient sample size to explore associations by race/ethnicity, and status of these perceptions during a later stage of the COVID-19 pandemic.

This study had limitations. Selection bias associated with online surveys is well established, for example, underrepresenting individuals who are older, without internet access, have lower income, and have less formal education; this effect is difficult to quantify, in either direction or magnitude, and may limit the generalizability of our results. However, the digital divide in internet access has shrunk over time.^25^ Despite our large sample size, samples for strata of important participant characteristics, including certain racial and ethnic minorities, were too small to provide sufficient statistical power for our analyses; still, we had sufficient statistical power to examine racial and ethnic differences between Black/African American, Hispanic/Latino, and White/Caucasian groups, which very few studies have done. Our questionnaire did not collect data on some characteristics that could affect risk perceptions, including presence of underlying health conditions, type of employment, or whether the respondent knew someone who had been infected with COVID-19.

## CONCLUSION

We found significant variations in perceived risk of COVID-19 transmission for social interaction and essential activities by age, gender, political affiliation, and race/ethnicity. These findings suggest the importance of socioeconomic differences, health disparities, and structural racism for efforts to control the COVID-19 pandemic, including preventive behaviors, care seeking for testing and treatment, and vaccination strategies. Further research should address how evidence-based interventions and programs can be tailored in consideration of these barriers with a goal of increased health equity in the pandemic response.

## Data Availability

Data can be made available upon request.

## Acknowledgments

We appreciate the team at Dynata for working closely with us during collection of the data. We would also like to recognize the Johns Hopkins University COVID-19 Research Response Fund for their initial support in getting this project off the ground. Thank you also to Dr. Gregory Kirk for help in developing the initial project plan. Lastly, thank you to the Johnson & Johnson Foundation for supporting this research project.

## Data availability

Data can be made available upon request.

## Authors’ contribution

SM, SS, DG, SA, and AL created the questionnaire and designed the survey. DG worked with Dynata to collect the data. DE, AZ, and PB conducted the analysis and drafted the manuscript. All authors contributed to the analysis, interpretation of the results, and reviewed and provided inputs to the manuscript.

**Supplementary Table 1:**
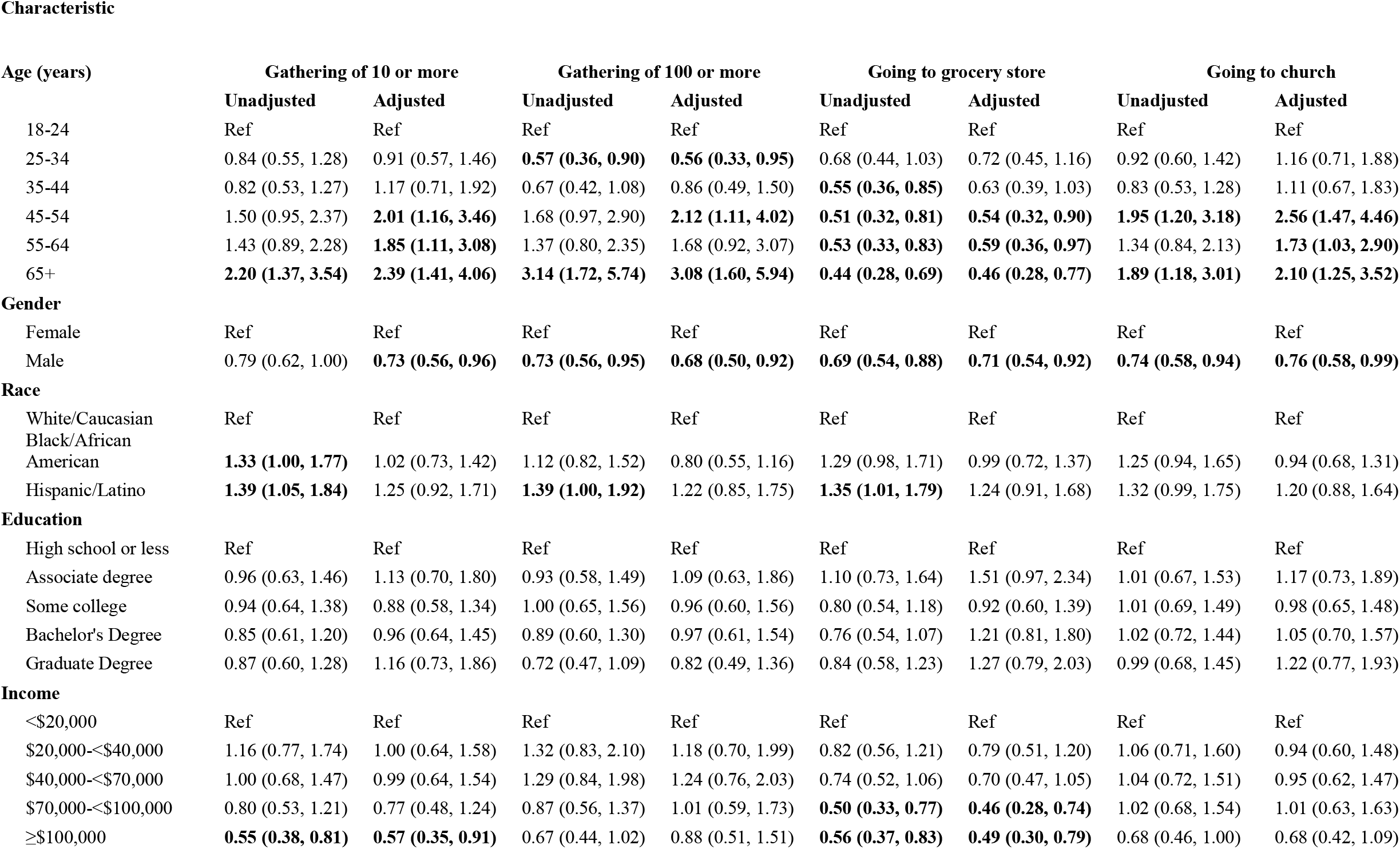
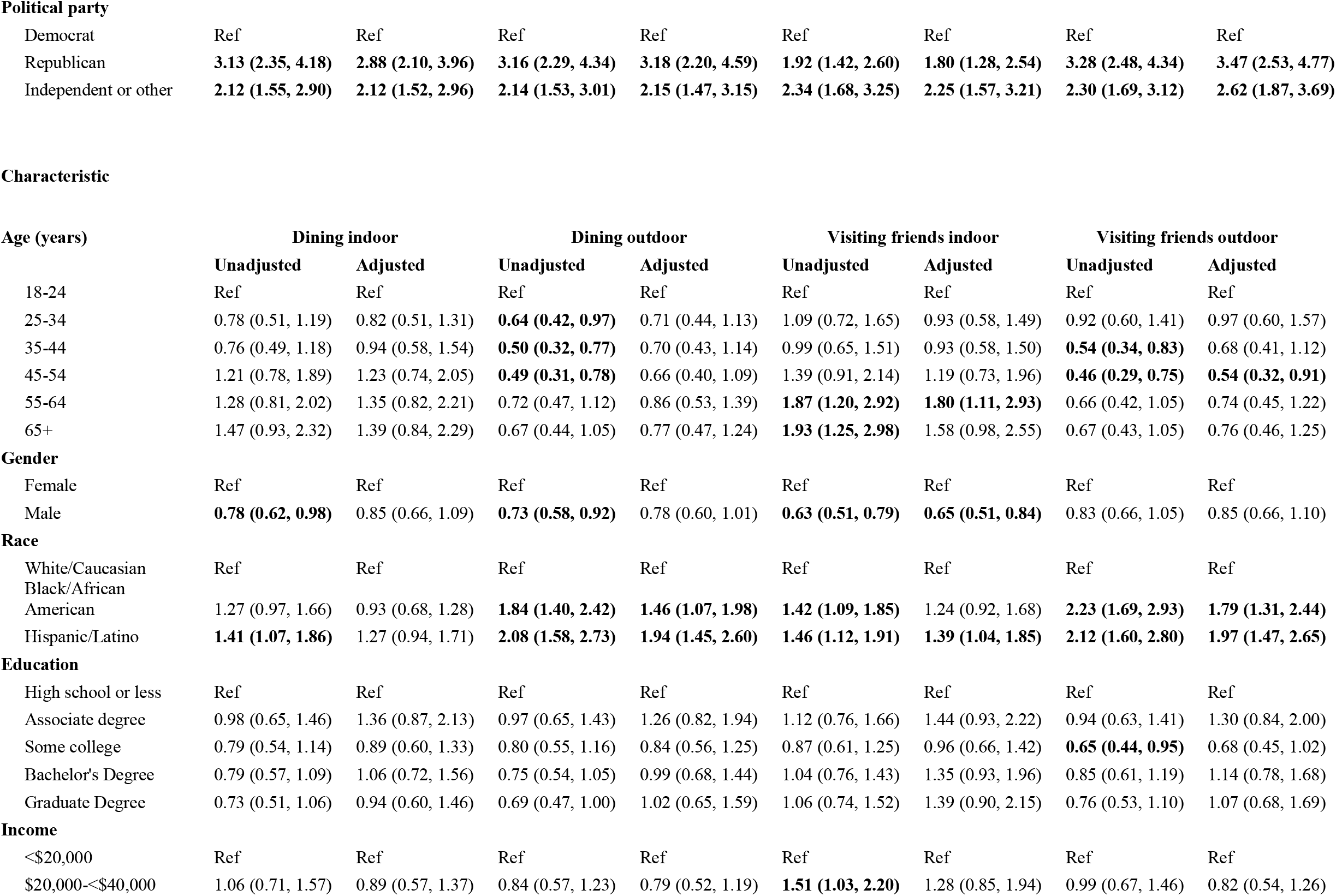
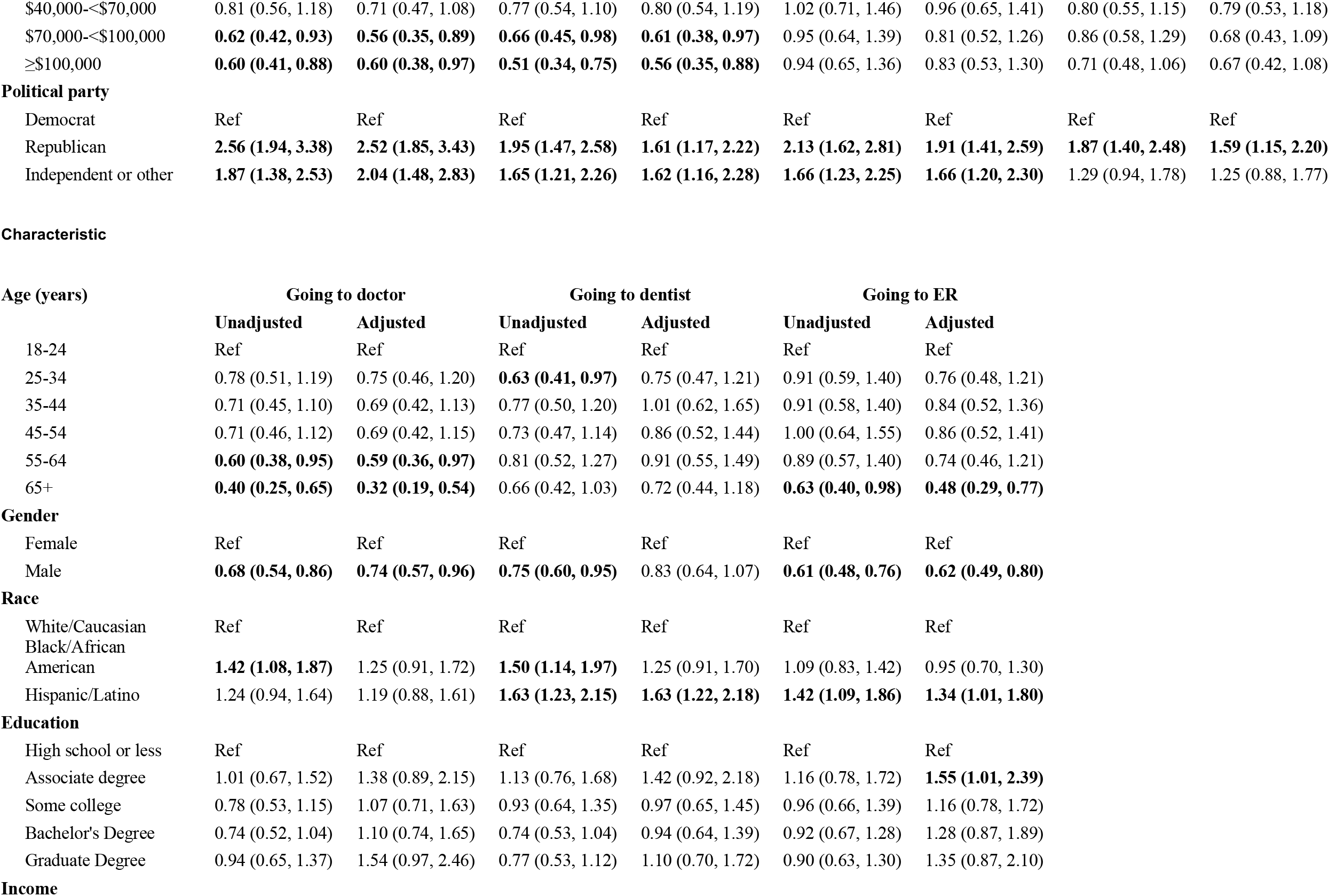
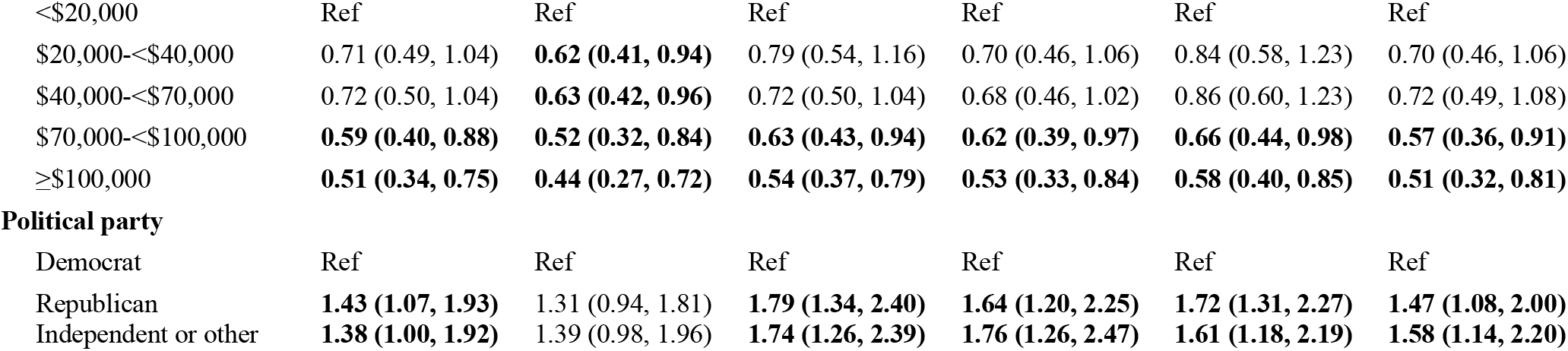
Unadjusted and adjusted Odds Ratios and 95% CIs for Perceiving Activities as Unsafe.

**Supplementary Table 2:**
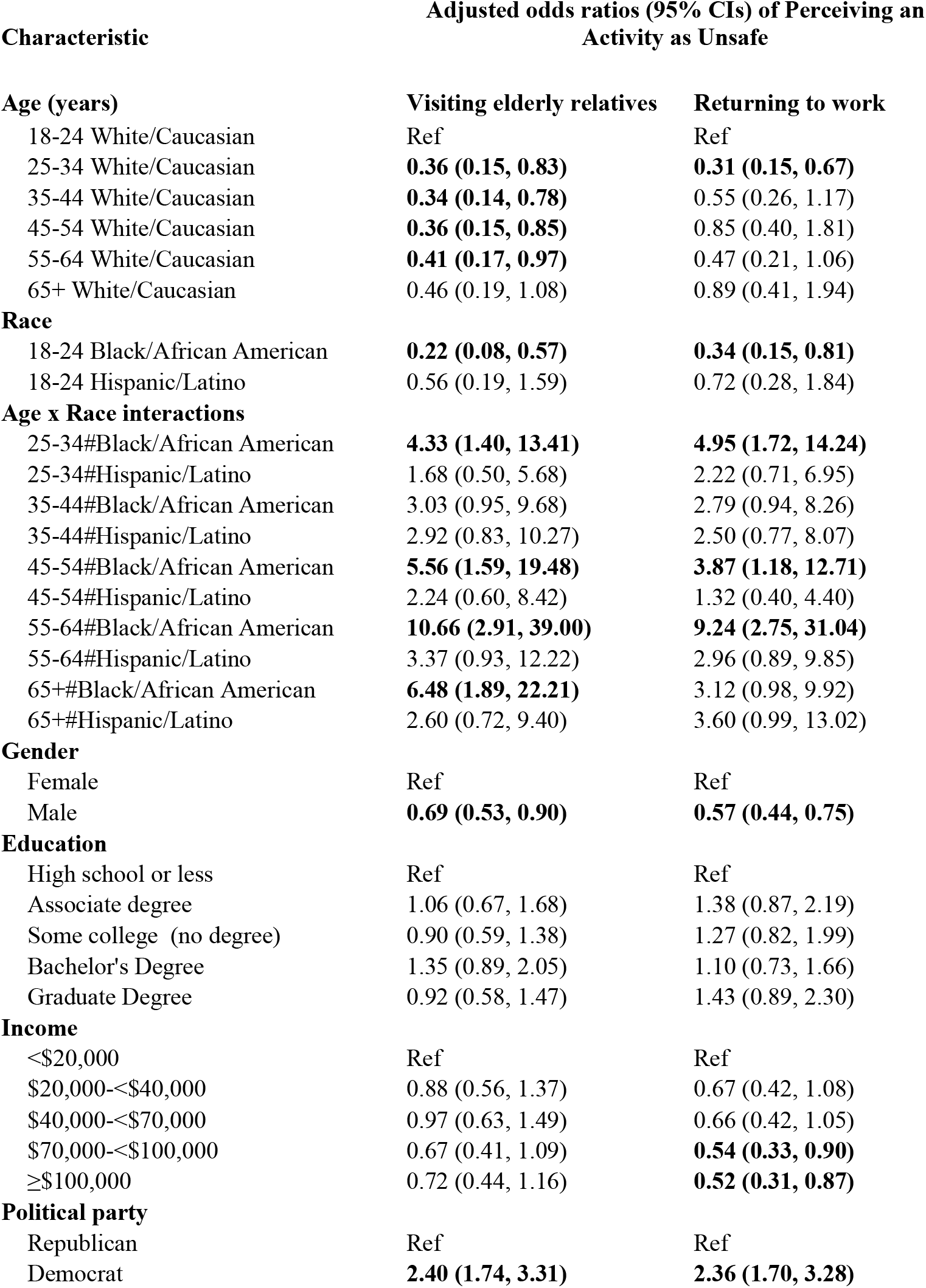
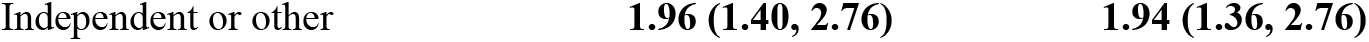
Adjusted Odds Ratios and 95% CIs for Perceiving Visiting Elderly Relatives and Returning to Work as Unsafe with Interaction Term for Age and Race.

**Supplementary Figure 1:**
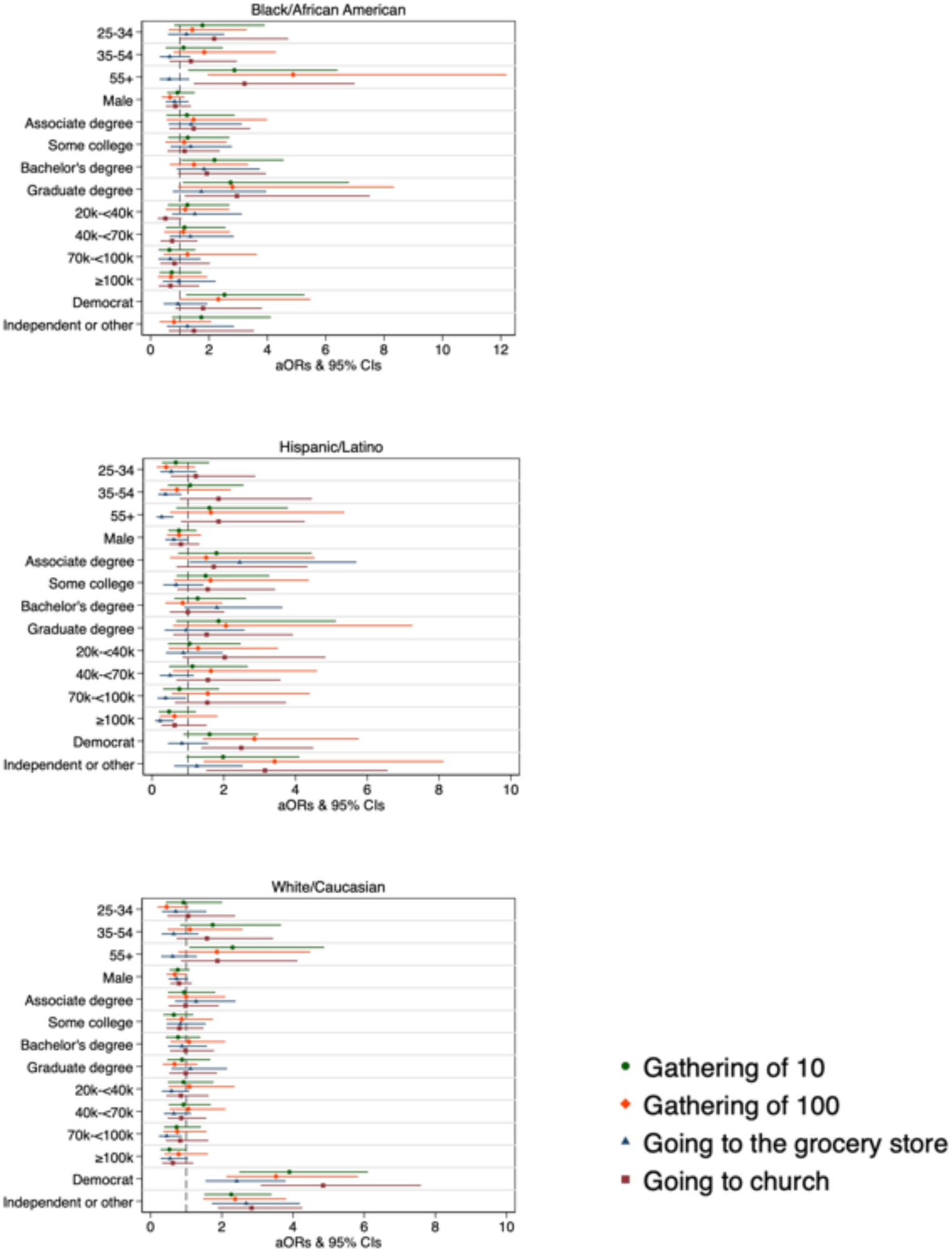
Adjusted odds ratios of perceiving large gatherings and activities in public as unsafe for participants stratified by race.

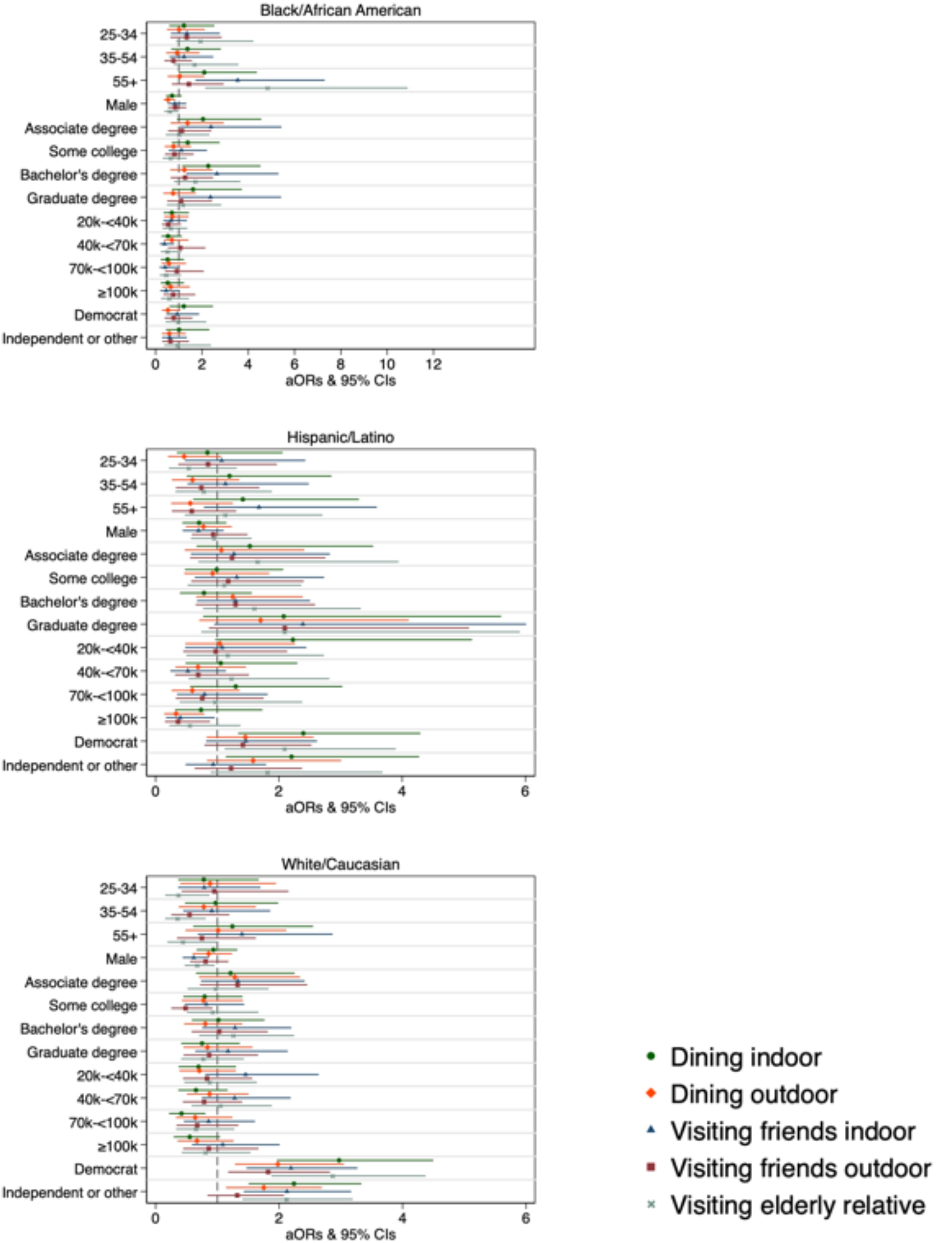
Adjusted odds ratios of perceiving indoor and outdoor dining and visits with friends and relatives as unsafe participants stratified by race.

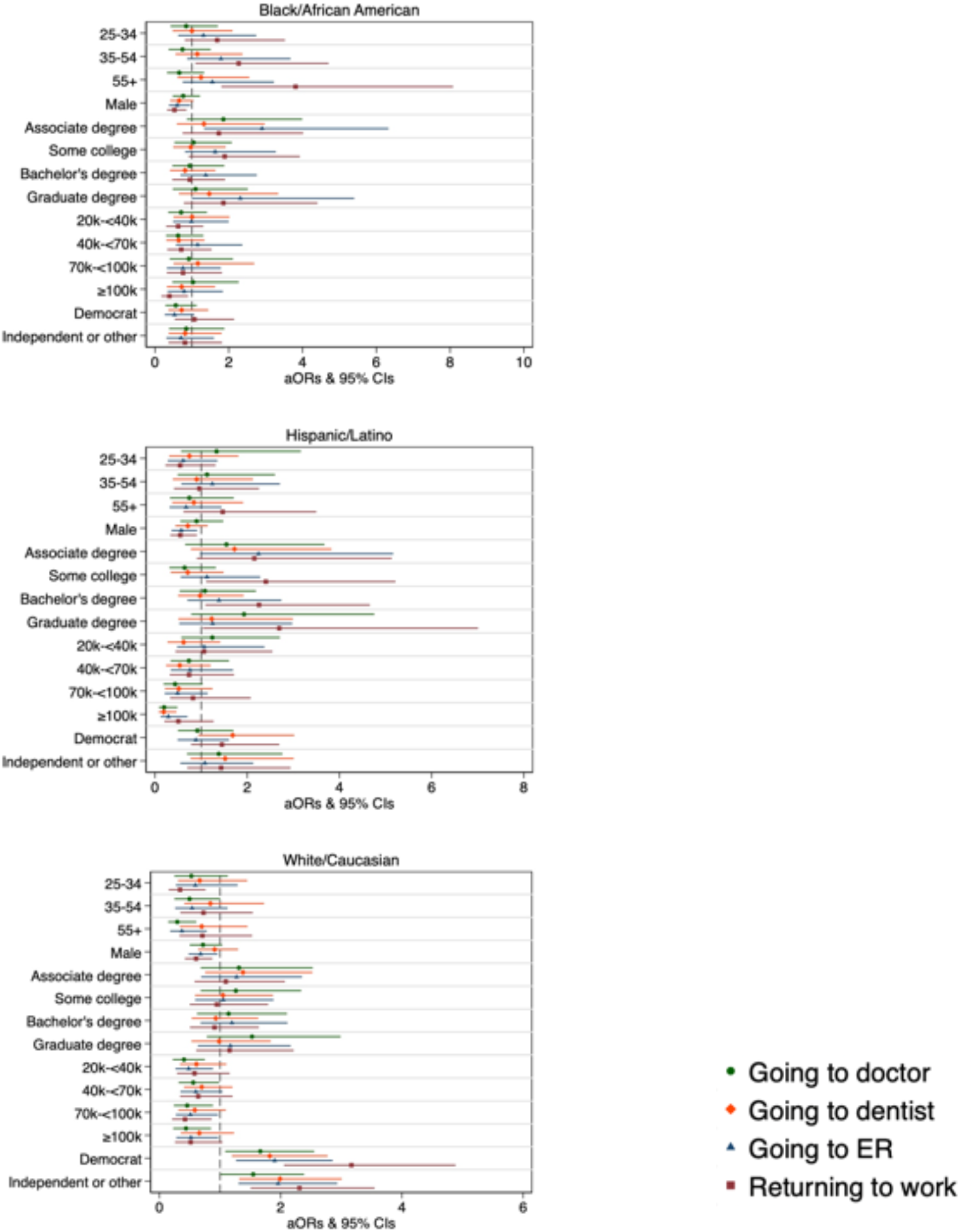
Adjusted odds ratios of perceiving medical visits and returning to work as unsafe for participants stratified by race.

